# Naturalistic visual search captures attentional disruption in mild cognitive impairment across distractor conditions

**DOI:** 10.64898/2026.09.08.26355011

**Authors:** Michael J. Kleiman, James E. Galvin

**Author notes:** Corresponding Author: M. J. Kleiman, Comprehensive Center for Brain Health, Department of Neurology University of Miami Miller School of Medicine, 7700 West Camino Real, Suite 200 Boca Raton, FL, USA.

## Abstract

Early detection of mild cognitive impairment (MCI) can be reliably achieved using measures of executive functions, however executive compensation can mask these changes in the earliest stages. Gaze behavior, specifically during visual search, has been hypothesized to measure executive functions and attention within MCI, but prior study has been inconclusive. This study characterized naturalistic visual search in MCI across distractor conditions and evaluated multivariate gaze-based classification of MCI from cognitively normal aging. In a cross-sectional, propensity-matched cohort of 62 CN and 62 MCI participants, participants completed a naturalistic scene-viewing search task under five distractor conditions (none, dissimilar, similar, background, and unexpected). Significant interactions were observed including in target dwell ratio and scanpath entropy, with trial-to-trial variability in area-of-interest coverage found to be higher in MCI. Repeated 10×5-fold cross-validation with feature selection discriminated CN from MCI with an area under the curve of 0.732 (95% CI [0.645, 0.818]); an expert-curated 14-feature panel reached 0.898 (95% CI [0.840, 0.949]). MCI-associated search disruption is displayed across qualitatively distinct distractor contexts, including in the absence of distractors, suggesting that naturalistic visual search merits evaluation as a candidate early-detection measure.

## Introduction

Early detection of mild cognitive impairment (MCI) has become a public-health imperative as the window for meaningful intervention continues to narrow [1,2]. **Neurobehavioral markers**, a category of digital biomarkers that index neurological or cognitive change through behaviors such as gaze, speech, and gait, have been proposed as an ecologically valid complement to conventional clinic-based assessment, capturing function as it is expressed during tasks that resemble real-world activity rather than during isolated cognitive assessments [3,4]. Established examples of this approach span several behavioral domains: connected speech analysis during picture description, [5,6] gait analysis under single- and dual-task conditions [7,8], and virtual reproduction of activities of daily living [9] each exhibit an ability to detect prodromal cognitive impairment. Gaze behavior in particular has a strong potential for capturing such markers of impairment [3,10,11], as eye-tracking is both passive and non-invasive, and captures the allocation of attention without necessarily requiring explicit instructions or verbal responses. Visual search is an especially informative domain for this purpose: because it requires active attentional selection among competing elements, it engages precisely the attentional processes that are vulnerable in MCI [10,12], making it a natural setting in which to look for gaze-based markers of impairment.

Eye-tracking studies have documented oculomotor search differences between MCI and cognitively normal (CN) adults across fixation count, dwell time, and distractor engagement parameters [13–16]. The systematic review by Wolf et al. [15] established the scope of this literature and identified a consistent limitation running through it: the paradigms are predominantly research tasks, with ecologically valid settings that engage the attentional demands of everyday visual environments underrepresented. Among the few studies that have examined real-world scenes, aggregate scanning metrics collapsed across trial structure have been the norm, with no significant differences found between amnestic MCI and controls at the agg[17]aslanboz2023]. The unresolved question is therefore whether MCI-associated search disruption is genuinely absent under naturalistic conditions or instead masked by collapsing across distinct contexts.

Each of the prior studies bearing on this question, however, leaves part of it unaddressed. Xue et al. [13] and Belan et al. [14] relied on single-paradigm laboratory tasks (noise-masking and eye-tracking-assisted visual inference language tasks, respectively) without a naturalistic protocol spanning qualitatively distinct types. Wolf et al. [15] explicitly called for ecologically valid settings yet found no prior study comparing MCI search across multiple distractor types within a single naturalistic protocol. Eraslan Boz et al. [17] used real-world scenes but analyzed only aggregate metrics, with neither a distractor-type comparison nor propensity matching to control for the age- and education-related differences that themselves shape search behavior. Yamada et al. [18], Camargo et al. [19], and Akkoyun et al. [10] compared MCI and CN search performance but, again, without a multi-distractor-type within-protocol comparison. This study addresses these gaps through three corresponding design elements: a naturalistic protocol spanning five qualitatively distinct distractor conditions; propensity matching to equate the groups on age, education, and sex; and a within-protocol comparison that characterizes the pattern of disruption across distractor types simultaneously. A further gap concerns classification: prior work has not evaluated whether gaze features drawn from a naturalistic multi-distractor protocol support multivariate discrimination of MCI from CN aging, and establishing which search behaviors carry that discrimination would clarify what a naturalistic search paradigm measures at the prodromal stage.

Here, we first test whether MCI-associated search disruption varies across qualitatively distinct distractor conditions; given the prior aggregate-null finding, condition-specific expression was expected. Second, we identify which distractor conditions produce the largest CN-vs-MCI effects, and which gaze features carry them. Next, we evaluate the multivariate classification accuracy of the gaze feature set for distinguishing MCI from CN aging. We further evaluate whether this discrimination and the univariate group differences generalize to an unmatched sample retaining all eligible participants. Finally, we characterize classification and univariate profiles separately for MCI attributable to Alzheimer’s disease (MCI-AD) and for MCI of other or mixed etiology (MCI-Other). The analyses revealed that search disruption was expressed across distractor types in a pattern that was largely attenuated at the whole-task aggregate level, and that a multivariate gaze classifier with curated feature selection discriminated MCI from CN aging at a high level of accuracy.

## Methods

### Participants

Participants were drawn from the Healthy Brain Initiative (HBI), a longitudinal research cohort of the Comprehensive Center for Brain Health [20]. All study procedures were approved by the University of Miami IRB, and written informed consent was obtained from every participant prior to enrollment. All participants underwent screening for neurological and psychiatric diagnoses that would be expected to confound visual or oculomotor performance. Participants were asked whether they normally wear glasses or contact lenses for reading or computer use, and any participant who reported doing so but was not wearing the correction at testing was excluded from analysis. No a priori power analysis was conducted; a post-hoc power analysis for the primary matched CN-vs-MCI comparison (N = 62 per group, α = .05, two-tailed) indicated 96% power to detect an effect of the magnitude of the largest false-discovery-rate-surviving condition-specific effect observed in this study (distractor dwell ratio in the perceptually similar condition, d = 0.671), and 58% power to detect an effect of the magnitude of the smallest surviving effect (target dwell ratio in the perceptually similar condition, d = 0.390).

Participants were classified according to the diagnostic criteria specified in the HBI protocol [20]. Within the MCI group, etiology was assigned for each participant by detailed consensus diagnosis incorporating neurological, cognitive, physical, and functional assessment with MRI and plasma biomarkers, including plasma pTau217, pTau181, and the Aβ42/40 ratio. This procedure yielded two etiology-defined subtypes: MCI-AD (N = 34) and MCI-Other (N = 28), the latter comprising cases attributed to dementia with Lewy bodies (DLB), vascular contributions (VCID), and mixed or other etiologies including Parkinson’s disease (PD), but not including non-neurodegenerative causes such as depression or normal-pressure encephalopathy.

Two analytic samples were constructed from this pool. The primary sample matched a CN sample to the 62 MCI participants on age, sex, and years of education. Matching was carried out among the eligible control participants who reported no cognitive concerns and produced matched groups of 62 CN and 62 MCI participants. The second sample was a held-out generalization set comprising the 98 control participants who reported no cognitive concerns and were not selected for matching, together with the same 62 MCI participants, so that no control contributed to both analyses. Because the diagnostic groups in this sample are not equated by construction and differ on age, sex, and education, classifier analyses conducted on it residualized those three variables from the gaze measures within each training fold rather than removing the imbalance by matching, so that no information from a held-out fold entered the adjustment.

### Materials and Measures

The visual search task analyzed here was administered as one component of the BACAN multimodal cognitive battery, a research platform that delivers a sequence of cognitive tasks within a single session; only the visual search task is considered in the present analysis. Eye movements were recorded with a Tobii Pro Spectrum eye tracker (Tobii AB, Danderyd, Sweden) operating at a sampling rate of 300 Hz. The search task presented naturalistic scenes under five qualitatively distinct distractor conditions: a no-distractor baseline condition; a condition with perceptually dissimilar distractors (e.g., cows and birds); a condition with perceptually similar distractors (e.g., cows and sheep); a condition in which the background itself constituted an interesting distractor; and a condition containing strange or unexpected distractors (e.g., fish in the sky).

The measurement modalities entering the present analysis comprised gaze-based features, including fixation counts, dwell times, scanpath entropy, area-of-interest coverage proportion, and distractor revisit count. Target and distractor regions of interest (AOIs) were drawn manually by the principal investigator (MJK) for each stimulus across all five distractor conditions, prior to data collection. After each search trial, participants were asked to report the number of targets present via a numeric keypress on a five-key subset of the keyboard’s numeric keypad, mapped to an on-screen response form whose key-to-count layout varied across trials so that no single key was consistently associated with a given count. Response accuracy was scored as whether this keypress matched the trial’s predefined correct count, and response delay was measured as the latency to the keypress, with a 30-second response window.

### Procedure

The present analysis constituted a cross-sectional observational analysis of performance at a single time point, drawing on data collected during a single research visit within the HBI longitudinal cohort study. After two no-distractor and two dissimilar-distractor trials, presentation order was randomized with a minimum of two trials presented for each distractor type. Participants were seated at a viewing distance from the stimulus display determined by Tobii Eye Tracker Manager, an application that guided the experimenter to position each participant at an optimal angle and distance from the tracker; the eye tracker was calibrated for each participant prior to the search task using a five-point calibration procedure, with recordings accepted only when both a visual assessment of calibration quality and the application’s automatic calibration failure detection confirmed adequate calibration.

Fixations were identified from the raw gaze stream using a velocity-threshold (I-VT) algorithm: a five-sample rolling mean was applied to the gaze samples prior to classification, samples with a velocity at or below 2.55 px/ms (corresponding to 30°/s) were classified as belonging to fixations and samples above this threshold as belonging to saccades, and no dispersion threshold was applied; fixations shorter than 50 ms were labeled as short fixations and were subject to merging. From the resulting gaze data, an extensive candidate feature pool of approximately 17,569 search-task-specific features was derived through a spatially dense grid-based decomposition of the stimulus screen; response-timing features were excluded from this classifier feature pool, though response accuracy and response delay were retained as dependent variables in the condition-structured analyses of variance. AOI-based ratio features were defined as the proportion of total trial dwell or fixation count attributable to a given region, comprising target dwell ratio, target fixation count ratio, distractor dwell ratio, and distractor fixation count ratio. Two AOI-based entropy measures were computed over the named AOIs rather than over a spatial grid: scanpath entropy, defined as the standard Shannon entropy H = −Σ p·log₂(p), in bits, over the proportion of fixations falling in each named AOI excluding the background; and transition-matrix entropy, defined as the weighted sum of the per-source-AOI row entropies of the AOI-to-AOI transition matrix, with each source AOI’s row entropy weighted by that AOI’s share of all transitions. Beyond the AOI-based ratio and entropy measures defined above, four further oculomotor measures reported in the Results were derived from the same fixation-and-saccade classification described earlier in this subsection.

Total saccade distance was defined as the summed length of every saccade identified within a trial. Mean saccadic direction change was defined as the average, across all saccades in a trial, of the absolute change in heading between each saccade and the one immediately preceding it. Mean microsaccade amplitude was defined as the average amplitude of the subset of saccade events classified as microsaccades. Blink rate was computed as the number of blink events divided by the total gaze-tracking duration for the trial.

### Statistical Analysis

Two complementary mixed-design analyses of variance (ANOVAs) were used to characterize how search behavior differed across distractor conditions. The first was a three-condition analysis modeling a within-subject Disruption Level factor (baseline, similar, and strange) and a between-subject Group factor (CN vs MCI); the second was a five-category strategy-specific analysis modeling a within-subject Category factor with all five distractor conditions as levels and the same between-subject Group factor. Age, sex, and race-ethnicity were entered as covariates in both models and in the aggregate exploration analyses. Effect sizes were reported as partial η² for all analysis-of-variance tests and as Cohen’s d for pairwise post-hoc comparisons, computed as the CN mean minus the MCI mean, so that a positive value denotes a higher score in the CN group and a negative value a higher score in the MCI group; the direction of every reported effect is additionally stated in words. Missing data was handled by available-case analysis applied separately to each dependent variable: for each model, participants with a missing value on that dependent variable, the grouping factor, or a covariate were dropped, so the number of participants contributing and the denominator degrees of freedom vary across dependent variables. ANOVA p-values are reported uncorrected. False discovery rate control was applied through the Benjamini-Hochberg procedure to the pairwise post-hoc contrasts within dependent-variable families, each family comprising the two to three pairwise contrasts associated with a single dependent variable. All analyses were conducted in Python 3.10.11 using SciPy 1.15.3, Pingouin 0.5.5, statsmodels 0.14.5, and scikit-learn 1.7.1.

Multivariate classification used elastic-net logistic-regression models evaluated under repeated stratified k-fold cross-validation (five folds, ten repeats, with stratification on the binary CN-vs-MCI label). Feature selection was nested within the cross-validation loop: within each training fold, elastic-net logistic-regression classifiers were fit to the feature pool, SHapley Additive exPlanations (SHAP) values were computed, and features carrying non-zero SHAP contributions were retained for that fold’s model, with pruning for high missingness, low variance, and pairwise correlation exceeding |r| = 0.85 applied within the fold; because selection is re-derived in every fold, no single fixed feature set is produced. Ninety-five percent confidence intervals were obtained by bootstrap resampling with 1,000 replicates, and the chance baseline was established by refitting the full pipeline to shuffled diagnostic labels. Operating characteristics were reported at the Youden-index threshold, defined as the decision threshold maximizing the sum of sensitivity and specificity. Sensitivity of the discrimination estimate to the particular selection of matched CN participants was assessed by redrawing the match independently across 20 draws, each evaluated under a single five-fold cross-validation repeat. A second classifier was evaluated on a fixed 14-feature panel held constant across all folds and evaluated under five-fold cross-validation with twenty repeats and no within-fold feature selection. The panel was curated pre-analysis by the investigators to span distractor engagement, saccadic dynamics, fixation-duration variability, and the organization of transitions between scene regions. The full composition of this panel (the operational definition of each of the fourteen features and the distractor condition it is computed within) is provided in **Table 1**. The fourteen selected features were examined within the matched sample and in the generalization sample separately, with Benjamini-Hochberg false discovery rate correction within each sample. Subtype analyses applied analyses of covariance (ANCOVAs) with age, sex, and race-ethnicity as covariates over the fixed fourteen-feature panel, beginning with an omnibus one-way model contrasting the three groups (CN, MCI-AD, and MCI-Other) and followed by pairwise comparisons with false discovery rate correction. Because splitting the MCI group breaks the one-to-one match, both subtype classifiers were trained on covariate-residualized features rather than on the matched contrast.

**Table 1.** The 14-feature panel.

| **Feature** | **Functional category** | **Distractor condition** | **Operational definition** |
| --- | --- | --- | --- |
| Transition-matrix entropy | Scanpath organization | Aggregated across all search stimuli | Count-weighted Shannon entropy of AOI-to-AOI gaze-transition probabilities; higher values indicate less predictable transitions between scene regions. |
| Blink-rate variability | Attention/arousal | Aggregated across all search stimuli | Trial-to-trial coefficient of variation in blink rate. |
| Distractor fixation-duration variability | Distractor engagement | Aggregated across all search stimuli | Trial-to-trial coefficient of variation in mean fixation duration on distractor regions. |
| Early distractor-dwell variability | Distractor engagement | Aggregated (first 2 s of each trial) | Trial-to-trial coefficient of variation in the proportion of the first 2 s of viewing time spent on distractor regions. |
| Within-trial fixation-duration slope | Scanpath organization | No-distractors | Ordinary-least-squares slope of fixation duration over ordinal fixation position within a trial. |
| Peak saccadic velocity toward the target | Target engagement | No distractors | Mean peak velocity of saccades directed toward the target, restricted to trials with no distractors present. |
| Early target fixation-count ratio | Target engagement | No-distractors (first 2 s) | Proportion of fixations landing on the target within the first 2 s of the trial. |
| Saccade-amplitude variability | Search strategy | High-similarity distractor | Standard deviation of saccade amplitude within a trial. |
| Distractor revisit count | Unexpected-distractor capture | Strange/unexpected distractor | Number of fixations returning to a distractor region after the target had already been fixated once. |
| Mean microsaccade amplitude | Unexpected-distractor capture | Strange/unexpected distractor | Mean amplitude of microsaccade events. |
| Mean saccadic direction change | Scanpath organization | Strange/unexpected distractor | Mean absolute change in saccade direction (degrees) between consecutive saccades. |
| First fixation on a distractor | Distractor engagement | Strange/unexpected distractor | Binary indicator of whether the first fixation of the trial landed on a distractor region. |
| First background-visit duration | Distractor engagement | Interesting-background-as-distractor | Duration of the first visit to the background region on trials with an interesting, distracting background |
| Background fixation duration | Distractor engagement | Interesting-background-as-distractor | Total fixation duration on the background region. |

## Results

### Sample Characteristics

Out of 503 total participants, 222 were assigned a diagnosis of CN or MCI (145 excluded as subjective-cognitive-impairment, 136 excluded as other diagnosis or poor data quality); propensity matching of the 160 CN participants to the 62 MCI participants on age, sex, and education yielded the final matched analytic sample reported below. Exclusions for uncorrected vision and for calibration or gaze-data-quality failure are applied upstream of the analysis pipeline during BACAN data collection and quality control and are not separately quantified in the analytic outputs available for this study. **Table 2** presents the demographic and clinical characteristics of the matched CN (CN, N = 62) and MCI (N = 62) groups, with the MCI sample further partitioned into etiology-defined subtypes of MCI-AD (N = 34) and MCI of other or mixed etiology (MCI-Other, N = 28). The CN group had a mean age of 72.27 years (SD = 7.88), was 51.61% female, and had completed a mean of 15.90 years of education (SD = 2.49). The MCI group had a mean age of 74.16 years (SD = 9.31), was 51.61% female, and had completed a mean of 15.27 years of education (SD = 2.88). Sex was an exact matching variable, so the two groups are identical in sex composition by construction, and age and education were equated by continuous nearest-neighbor matching. The MCI-AD and MCI-Other subtypes did not differ significantly on age (MCI-AD 75.82 ± 8.72; MCI-Other 72.14 ± 9.76 years; F(2,121) = 2.17, p = .118) or years of education (MCI-AD 15.12 ± 2.71; MCI-Other 15.46 ± 3.12; F(2,121) = 0.97, p = .384), and sex composition was likewise comparable across the three groups (58.82% female in MCI-AD, 42.86% in MCI-Other; χ²(2) = 1.57, p = .457). Age, sex, and race-ethnicity were retained as covariates in all main analyses. Cognitive and clinical severity differed between groups, as expected: the CN group scored higher than the MCI group on the MoCA (26.85 ± 2.25 vs. 22.74 ± 2.92; Welch’s t = 8.78, p < .001, d = 1.58), Number Symbol Coding (42.42 ± 9.06 vs. 31.84 ± 7.68; t = 7.02, p < .001, d = 1.26), the Resilience Index (186.01 ± 29.99 vs. 166.73 ± 32.85; t = 3.41, p < .001, d = 0.61), and the Brain Health Index (66.61 ± 8.90 vs. 56.55 ± 9.11; t = 6.22, p < .001, d = 1.12), and lower on the Vulnerability Index (7.32 ± 2.92 vs. 9.02 ± 3.02; t = −3.17, p = .002, d = −0.57) and the CDR-SB (0.04 ± 0.14 vs. 0.94 ± 0.77; t = −9.06, p < .001, d = −1.63).

**Table 2.** Sample characteristics.

| Characteristic | CN<br>(n = 62) | MCI<br>(n = 62) | MCI-AD<br>(n = 34) | MCI-Other<br>(n = 28) | p-value<br>(CN vs MCI) |
| --- | --- | --- | --- | --- | --- |
| Age | 72.27 (7.88) | 74.16 (9.31) | 75.82 (8.72) | 72.14 (9.76) | .261 |
| %Female | 51.61% | 51.61% | 58.82% | 42.86% | 1.000 |
| Education (years) | 15.90 (2.49) | 15.27 (2.88) | 15.12 (2.71) | 15.46 (3.12) | .240 |
| Race/Ethnicity: Hispanic White | 8.06% | 11.29% | 14.71% | 7.14% | .598 |
| Race/Ethnicity: Non-Hispanic White | 82.26% | 67.74% | 73.53% | 60.71% | .080<sup>c</sup> |
| Race/Ethnicity: Non-Hispanic Black | 3.23% | 14.52% | 8.82% | 21.43% | .040<sup>c</sup> |
| Race/Ethnicity: Other | 6.45% | 6.45% | 2.94% | 10.71% | 1.000 |
| MoCA | 26.85 (2.25) | 22.74 (2.92) | 22.50 (2.76) | 23.04 (3.14) | < .001<sup>bc</sup> |
| Number Symbol Coding | 42.42 (9.06) | 31.84 (7.68) | 32.68 (7.35) | 30.82 (8.08) | < .001<sup>bc</sup> |
| Resilience Index | 186.01 (29.99) | 166.73 (32.85) | 172.16 (34.40) | 160.14 (30.16) | .002<sup>c</sup> |
| Vulnerability Index | 7.32 (2.92) | 9.02 (3.02) | 8.32 (2.84) | 9.86 (3.08) | .003<sup>ac</sup> |
| Brain Health Index | 66.61 (8.90) | 56.55 (9.11) | 58.50 (9.02) | 54.18 (8.79) | < .001<sup>bc</sup> |
| CDR Sum of Boxes | 0.04 (0.14) | 0.94 (0.77) | 0.75 (0.46) | 1.16 (0.98) | < .001<sup>abc</sup> |
| Trail Making Test A (s) | 30.68 (9.67) | 43.38 (18.29) | 38.44 (13.77) | 49.36 (21.36) | < .001<sup>abc</sup> |
| Trail Making Test B (s) | 71.26 (25.65) | 115.00 (44.49) | 105.12 (40.73) | 125.97 (46.64) | < .001<sup>bc</sup> |
| Verbal Fluency (animals) | 21.55 (3.78) | 17.47 (4.70) | 17.88 (4.47) | 16.96 (4.99) | < .001<sup>bc</sup> |
| Functional Activities Questionnaire | 0.39 (1.11) | 2.24 (4.16) | 1.18 (1.91) | 3.54 (5.61) | .002<sup>abc</sup> |
| Quick Dementia Rating Scale | 0.47 (0.77) | 2.02 (2.39) | 1.43 (1.89) | 2.75 (2.75) | < .001<sup>abc</sup> |
| Benson Figure Copy | 16.24 (1.29) | 15.68 (1.61) | 15.62 (1.48) | 15.75 (1.78) | .045<sup>b</sup> |
| Benson Figure Recall | 11.63 (2.93) | 8.03 (3.74) | 7.76 (4.04) | 8.36 (3.39) | < .001<sup>bc</sup> |
| HVLT Immediate Recall | 24.79 (4.03) | 17.89 (4.01) | 17.97 (4.20) | 17.79 (3.84) | < .001<sup>bc</sup> |
| HVLT Delayed Recall | 9.27 (1.82) | 4.95 (2.93) | 4.56 (2.90) | 5.43 (2.94) | < .001<sup>bc</sup> |

Note. M (SD) = mean (standard deviation). The MCI sample is divided into MCI due to Alzheimer's disease (MCI-AD, n = 34) and MCI of other or mixed etiology (MCI-Other, n = 28). The p-value column reports the overall CN vs. MCI comparison (one-way ANOVA for continuous measures, Pearson chi-square for categorical measures), false-discovery-rate corrected (Benjamini-Hochberg) across all rows. Superscript letters on the p column mark additional pairwise comparisons significant at $p < .05$ , uncorrected (Welch's t for continuous measures, Fisher's exact test for categorical measures): <sup>a</sup> MCI-AD vs. MCI-Other; <sup>b</sup> CN vs. MCI-AD; <sup>c</sup> CN vs. MCI-Other.

The generalization sample comprised a held-out set of 98 control participants who reported no cognitive concerns and were not selected for matching. Relative to the matched control group, this held-out control group was younger (mean age 67.05 years, SD = 8.30), more predominantly female (85.71%), and more educated (mean 16.86 years, SD = 1.62).

### Group Differences Across Distractor Conditions

Significant group main effects were observed for scanpath entropy (F(1,108) = 11.72, p < .001, partial η² = 0.098) and for target dwell ratio (F(1,106) = 9.53, p = .003, partial η² = 0.083), indicating lower overall target engagement and a narrower distribution of fixations across named regions in the MCI group across distractor conditions (**Figure 1**). Additional significant group main effects were observed for distractor dwell ratio (F(1,98) = 10.55, p = .002, partial η² = 0.097) and distractor revisit count (F(1,113) = 8.43, p = .004, partial η² = 0.069), both likewise lower in the MCI group.

**Figure 1.**
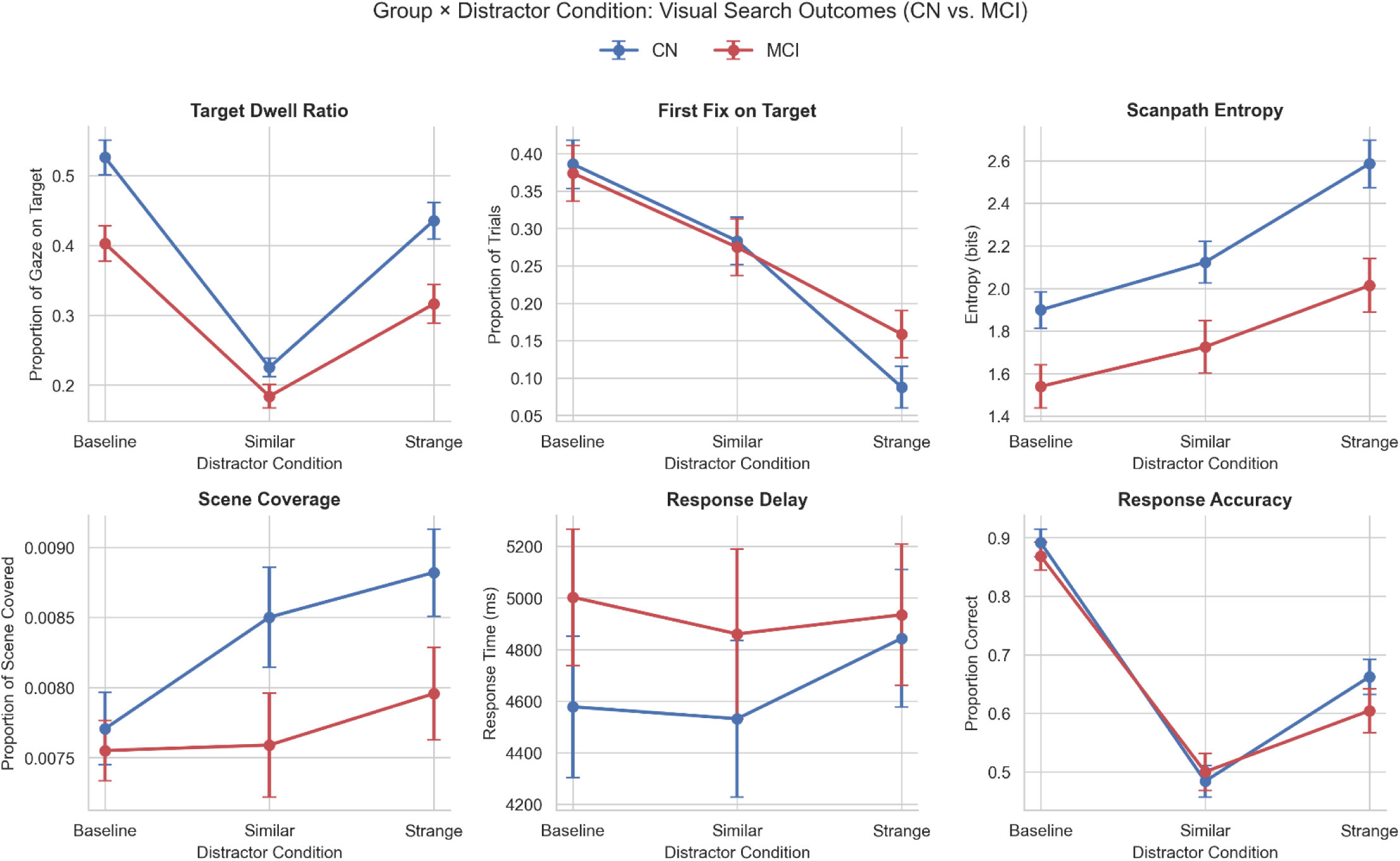
Group × distractor condition interaction plots for six visual search outcome measures in cognitively normal (CN; n = 62) and mild cognitive impairment (MCI; n = 62) participants. Data points represent group means; error bars indicate ±1 standard error of the mean. Distractor conditions correspond to experimental categories in the BACAN visual search task: Baseline, Similar, and Strange. Panel rows reflect two broad functional domains: target and search engagement (top; Target Dwell Ratio, First Fix on Target, Scanpath Entropy) and behavioral performance (bottom; Scene Coverage, Response Delay, Response Accuracy). Mixed-model ANOVAs covaried for age, sex, and years of education; pairwise post-hoc comparisons were FDR-corrected within each dependent variable.

The three-condition analysis yielded significant interactions for six of the 15 dependent variables tested: response accuracy (F(2,242) = 4.58, p = .011, partial η² = 0.036), distractor dwell ratio (F(1,98) = 4.63, p = .034, partial η² = 0.045), distractor fixation count ratio (F(1,98) = 4.58, p = .035, partial η² = 0.045), target dwell ratio (F(2,212) = 3.30, p = .039, partial η² = 0.030), total saccade distance (F(2,234) = 3.10, p = .047, partial η² = 0.026), and scanpath entropy (F(2,216) = 3.07, p = .048, partial η² = 0.028). The distractor dwell ratio and distractor fixation count ratio terms reflect the group-by-condition contrast across the two distractor-present levels, similar and strange.

Post-hoc CN-vs-MCI comparisons at each disruption level, for the dependent variables carrying a significant interaction, showed reduced engagement with both the named target and distractor regions in the MCI group at every disruption level, together with a correspondingly greater share of dwell on background and peripheral regions. Of the 41 contrasts tested, 11 survived FDR correction. The CN group exceeded the MCI group on distractor dwell ratio in the similar condition (d = 0.671, p = .002) and on distractor fixation count ratio in the same condition (d = 0.496, p = .023); on distractor revisit count under strange distractors (d = 0.498, p = .017) and in the similar condition (d = 0.426, p = .023); on target fixation count ratio at baseline (d = 0.537, p = .013); on target dwell ratio at baseline (d = 0.435, p = .029), in the similar condition (d = 0.390, p = .043), and under strange distractors (d = 0.461, p = .029); and on scanpath entropy in the similar condition (d = 0.451, p = .029) and under strange distractors (d = 0.497, p = .026); and on response accuracy under strange distractors (d = 0.452, p = .041).

### Strategy-Specific Vulnerability Profile

The five-category strategy-specific analysis yielded significant Group × Category interactions for five of the 14 dependent variables tested: scanpath entropy (F(4,412) = 4.24, p = .002, partial η² = 0.040), target dwell ratio (F(4,372) = 3.15, p = .015, partial η² = 0.033), target fixation count ratio within the first 4 s (F(4,328) = 3.15, p = .015, partial η² = 0.037), distractor revisit count (F(2,222) = 3.23, p = .042, partial η² = 0.028), and target fixation count ratio (F(4,372) = 2.57, p = .038, partial η² = 0.027), **Figure 2**.

**Figure 2.**
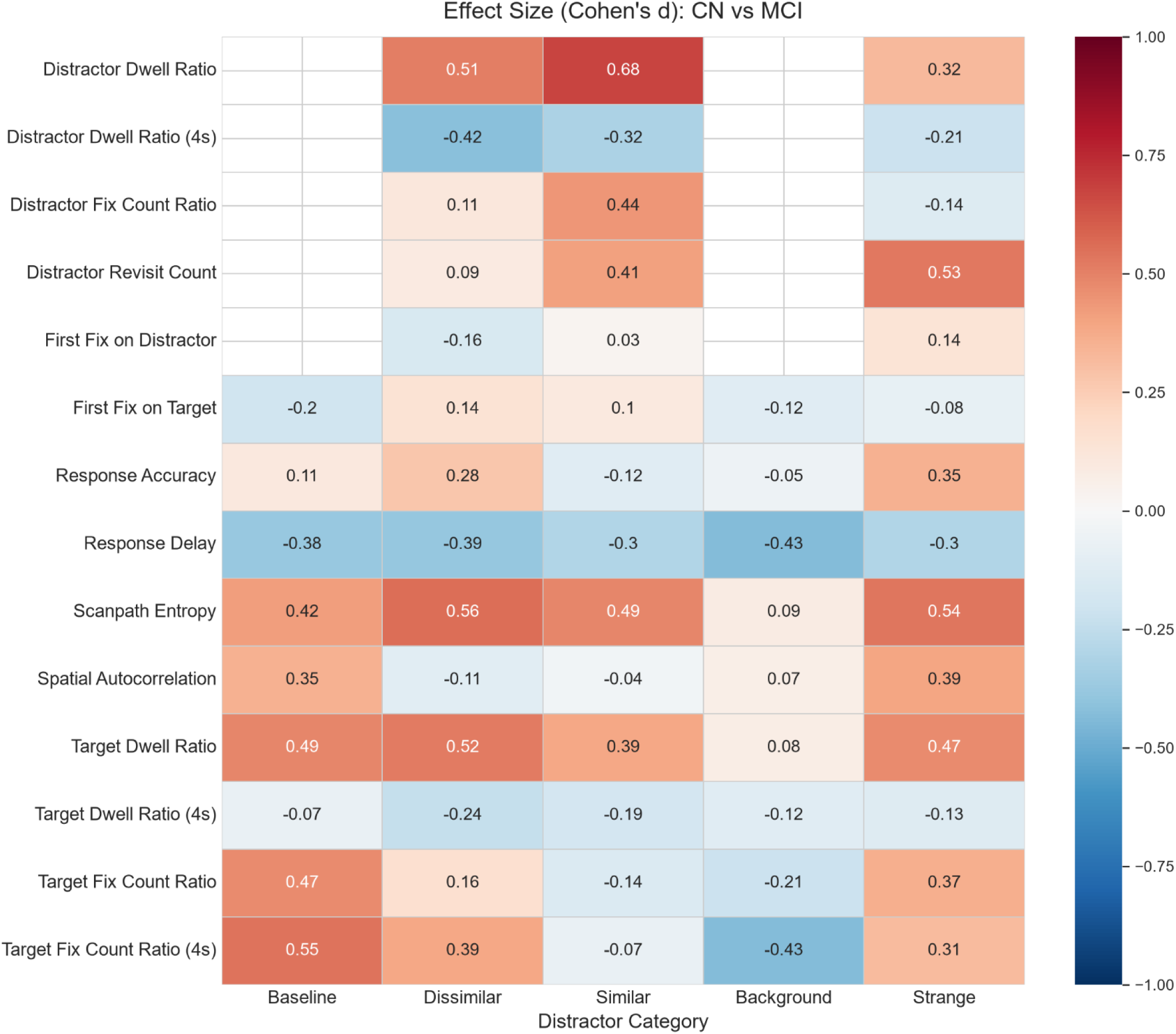
Cohen’s d effect size heatmap for CN vs. MCI group differences across gaze outcome measures at each distractor condition. Rows correspond to dependent variables; columns correspond to distractor category ordered by increasing cognitive disruption demand. Positive values (blue) indicate CN > MCI; negative values (red) indicate MCI > CN. Effect sizes were computed from covariate-adjusted pairwise comparisons following FDR correction within each dependent variable.

Decomposing these interactions into post-hoc effect sizes, the largest single CN-vs-MCI effect was distractor dwell ratio in the perceptually similar condition (d = 0.671, p = .002), followed by early target capture, indexed by target fixation count ratio within the first 4 s, at the no-distractor baseline (d = 0.597, p = .008), **Figure 3**. Of the 60 contrasts tested, 13 survived false discovery rate correction, each in the direction of a higher value in the CN group. Additional surviving effects included scanpath entropy under dissimilar distractors (d = 0.568, p = .016), target fixation count ratio at baseline (d = 0.537, p = .021), target dwell ratio under dissimilar distractors (d = 0.520, p = .032), distractor revisit count under strange distractors (d = 0.498, p = .025), scanpath entropy under strange distractors (d = 0.497, p = .022), distractor fixation count ratio in the similar condition (d = 0.496, p = .035), and distractor dwell ratio under dissimilar distractors (d = 0.494, p = .038). The distribution of effect sizes across conditions showed distractor engagement effects concentrated under perceptually similar distractors, target engagement effects concentrated at the no-distractor baseline, and scanpath entropy effects present under both dissimilar and strange distractors; effects under interesting-background distractors were smaller or non-significant.

**Figure 3.**
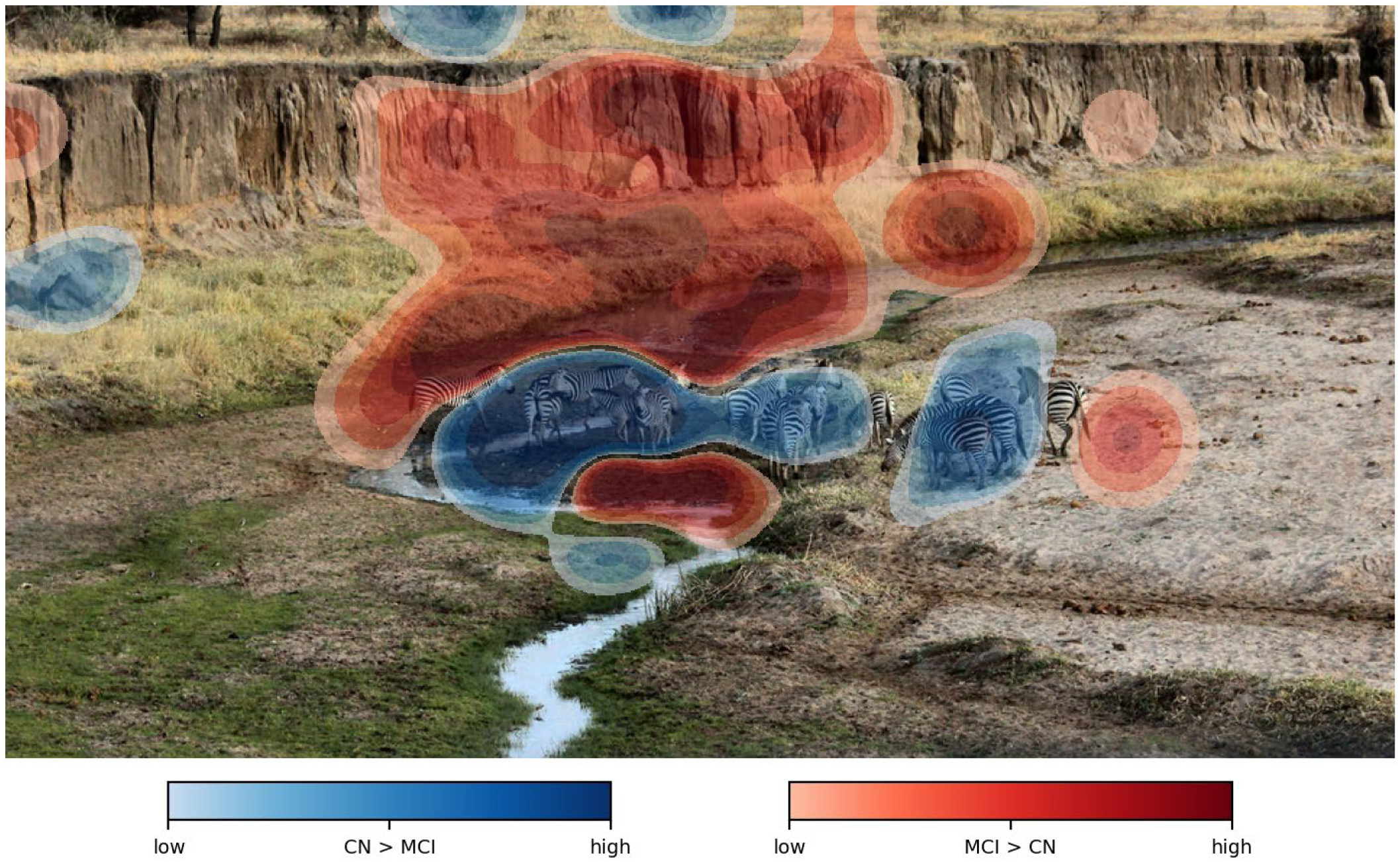
Early gaze difference map for the no-distractor condition, first 4 seconds. Fixation density was computed per subject and averaged separately for CN and MCI groups, then subtracted (CN − MCI). Densities are drawn from the 1:1 age-, sex-, and education-matched cohort (62 CN / 62 MCI). Blue regions indicate areas where CN subjects fixated more; red regions indicate areas where MCI subjects fixated more. In the no-distractor condition, CN subjects directed proportionally more early fixations to the target objects, while MCI subjects showed greater early dwell on the background and peripheral regions. This asymmetry corresponds to one of the strongest baseline search effects in the matched cohort: target fixation count ratio in the first 4 seconds at baseline (Cohen’s d = 0.55, p = .018).

### Aggregate Exploration Behavior

When gaze metrics were collapsed across distractor conditions, the group separation observed in the condition-structured analyses was largely, though not entirely, attenuated. Of the 25 aggregate exploration dependent variables tested, one survived false discovery rate correction: trial-to-trial variability in area-of-interest coverage was elevated in the MCI group (F = 11.00, p = .001, p = .030, partial η² = 0.086, d = −0.608). Five further measures reached the uncorrected threshold without surviving correction. Transition-matrix entropy was elevated in the MCI group (F = 7.67, p = .007, p = .082, d = −0.510), moving in the opposite direction to scanpath entropy, which was higher in the CN group (F = 4.45, p = .037, d = 0.384). Aggregate area-of-interest coverage proportion was likewise higher in the CN group at the uncorrected level (F = 5.40, p = .022, d = 0.423) but did not survive correction (p = .169). Variability in transition-matrix entropy was higher in the CN group (F = 4.46, p = .037, d = 0.396), and variability in early scene coverage was higher in the MCI group (F = 3.98, p = .048, d = −0.366). The single aggregate effect to survive correction was therefore a measure of exploratory inconsistency rather than a mean-level difference in exploratory behavior.

### Multivariate Classification

The full-feature elastic-net classifier, with feature selection nested within every cross-validation fold, discriminated CN from MCI with an area under the curve (AUC) of 0.732 (95% CI [0.645, 0.818]), retaining a mean of 72.4 features per fold from the pool of 17,569 search-task-specific candidate features; refitting the same pipeline to shuffled diagnostic labels yielded a mean AUC of 0.463, consistent with chance-level performance and suggesting that the observed discrimination is not an artifact of label leakage in the cross-validation procedure. At the Youden operating point (decision threshold 0.315), this classifier achieved a sensitivity of 0.887 (95% CI [0.797, 0.954]) and a specificity of 0.516 (95% CI [0.393, 0.638]). The classifier built on the fixed fourteen-feature panel discriminated CN from MCI with an AUC of 0.898 (95% CI [0.840, 0.949]), achieving a sensitivity of 0.968 (95% CI [0.914, 1.000]) and a specificity of 0.758 (95% CI [0.651, 0.860]).

The two classifiers were given different information at feature-selection time and address different questions of the same data; neither is a benchmark for the other. Sensitivity to the particular selection of matched CN participants was assessed with a 20-draw matching-stability check in which the match was redrawn independently on each draw. Across draws the full-feature model yielded a mean cross-validated AUC of 0.653 (SD = 0.030, range [0.577, 0.694]), below the value obtained on the primary matched sample. Each stability draw used a single cross-validation repeat against ten for the primary analysis, so the difference reflects both the design of the stability check and the particular matched sample drawn; the discrimination estimate should accordingly be read as specific to this matched sample rather than as a property of the paradigm.

The 14-feature panel, fitted under the same repeated cross-validation scheme but with age, sex, and education residualized from every measure within each training fold, was then applied to the held-out generalization sample of 98 cognitively normal controls with no cognitive concerns and the 62 MCI participants.

Discrimination in this sample reached an AUC of 0.701 (95% CI [0.625, 0.781]), with a sensitivity of 0.774 (95% CI [0.673, 0.881]) and a specificity of 0.612 (95% CI [0.510, 0.710]) at a decision threshold of 0.437.

### Feature-Level Effects

The 14 features composing the fixed panel were each tested for group differences one at a time, independently of any classifier, so that the behavioral content of the panel could be read without reference to how the measures combine. In the matched sample, 11 of the 14 measures separated the groups after false discovery rate correction; across the pool of search-task-specific gaze features profiled univariately, no individual feature’s discrimination exceeded an area under the curve of approximately 0.74. Participants with MCI returned to distractors less often under strange distractors, devoted a smaller share of early fixation to the target at the no-distractor baseline, dwelled less on the background in the condition in which the background itself served as the distractor, reached lower peak saccadic velocity on approach to the target, and produced less variable saccade amplitudes; they were also less variable in the proportion of early fixation duration directed at distractors. Moving in the opposite direction, participants with MCI showed greater trial-to-trial variability in mean distractor fixation duration and in blink rate, larger mean microsaccade amplitude under strange distractors, and higher transition-matrix entropy, indicating movement between scene regions that was less predictable from the region currently fixated.

In the generalization sample, 8 of the 14 measures separated the groups after correction. Seven of the fourteen were significant after correction in both samples, and each of those seven carried the same direction in each.

Half of the panel reproduces its group difference across the demographically matched sample and the held-out control group.

### MCI Subtype Profiles

Saccadic direction reversals (direction changes > 45°) during the strange-distractor task across three panels (CN, MCI-AD, and MCI-Other) illustrate elevated reversal density in the MCI-Other group relative to the CN and MCI-AD groups, **Figure 4**. The subtype analyses were conducted as ANCOVAs with age, sex, and race-ethnicity as covariates over the fixed fourteen-feature panel, comparing CN (N = 62), MCI-AD (N = 34), and MCI-Other (N = 28); both subtype cells exceed the minimum cell size prespecified for inference in this project, and MCI-Other is a heterogeneous grouping whose composition bounds the interpretation of every contrast it enters.

**Figure 4.**
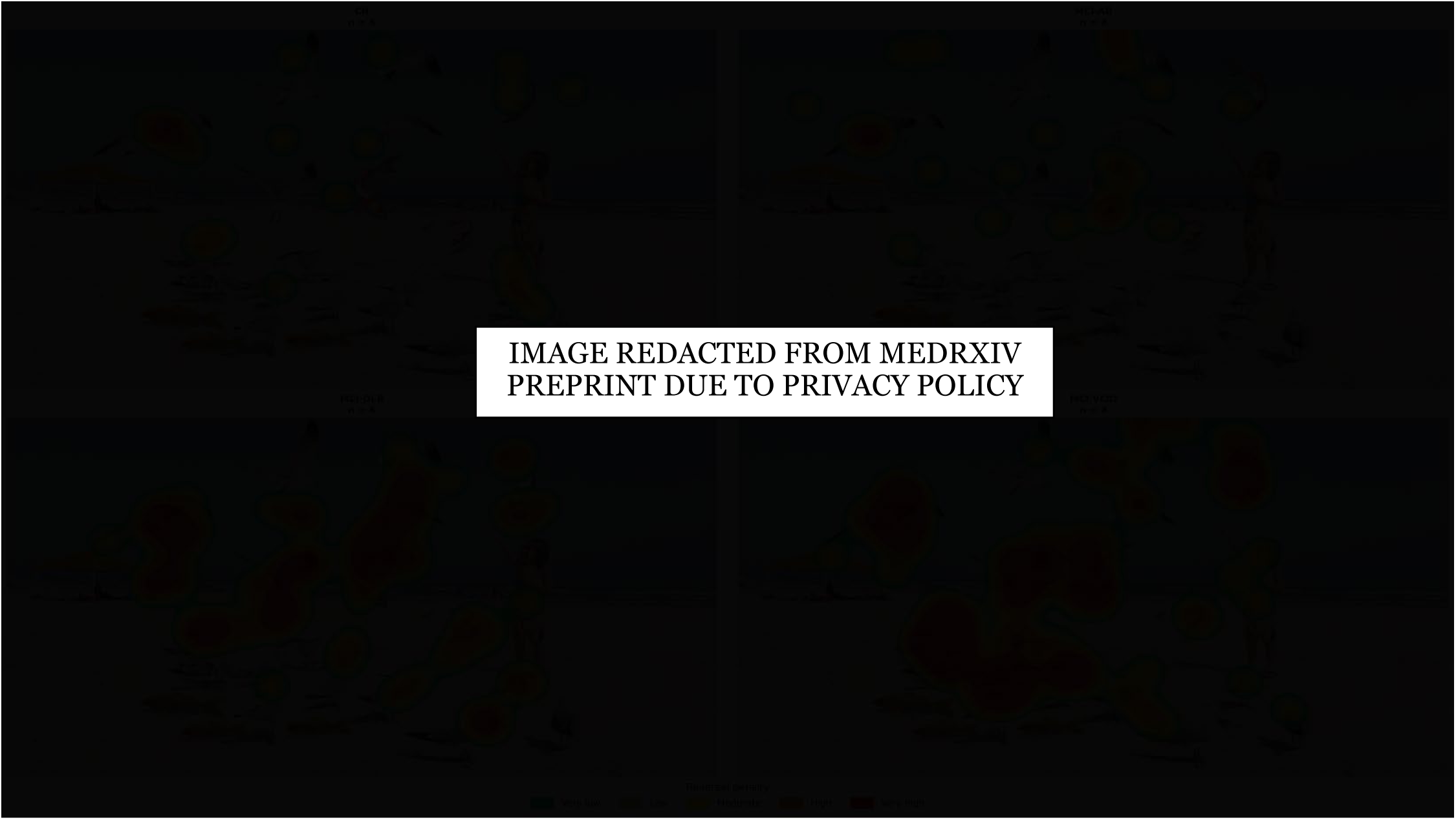
Spatial distribution of saccadic direction reversals during the strange-distractor visual search task (A110) across MCI subtypes. Reversal density maps showing the spatial locations of fixations that followed a saccadic direction change of ≥90°, for cognitively normal controls (CN), MCI due to Alzheimer’s disease (MCI-AD), MCI due to Lewy body disease (MCI-DLB), and MCI due to vascular disease (MCI-VCID), drawn from the 1:1 age-, sex-, and education-matched cohort (62 CN / 62 MCI). Each panel displays eight representative participants selected to illustrate the extremes of each group’s direction-change profile (CN and MCI-AD: participants with the fewest reversals; MCI-DLB and MCI-VCID: participants with the most reversals). Across all strange-distractor stimuli, mean saccadic direction change was significantly elevated in MCI groups relative to CN (one-way ANOVA: F(3, 114) = 4.96, p = .003), an effect that persisted after covarying for age, sex, and years of education (ANCOVA: F(3, 111) = 4.54, p = .005). Subgroup analysis revealed that this effect was driven specifically by the vascular and Lewy body subtypes: CN demonstrated significantly smaller direction changes than both MCI-DLB (p = .032, d = −1.06) and MCI-VCID (p = .032, d = −0.85), while MCI-AD did not differ significantly from CN (p = .074, d = −0.47). These results suggest that disordered saccadic control during naturalistic visual search is particularly pronounced in vascular and Lewy body MCI, consistent with the known involvement of posterior cortical and subcortical oculomotor circuits in these etiologies, and may offer a non-invasive behavioral signature for MCI subtype differentiation.

Across the subtype panel, 12 of the 14 features distinguished the three groups on the omnibus test after false discovery rate correction. The strongest omnibus effects were trial-to-trial variability in the proportion of early fixation duration devoted to distractors (F = 7.08, p = .017, partial η² = 0.120), transition-matrix entropy (F = 6.16, p = .017, partial η² = 0.097), and mean saccadic direction change under strange distractors (F = 5.87, p = .017, partial η² = 0.092); the two features not reaching significance were first background visit duration (p = .073) and whether the first fixation landed on a distractor under strange distractors (p = .454). Decomposing these into pairwise contrasts, five features separated CN participants from both MCI-AD and MCI-Other: summed background fixation duration in the interesting-background condition, distractor revisit count under strange distractors, two measures of distractor-fixation-duration variability, and peak saccadic velocity toward the target. Four features separated CN participants from MCI-Other only: blink-rate variability, microsaccade amplitude under strange distractors, transition-matrix entropy, and mean saccadic direction change under strange distractors, the last elevated in the MCI-Other group (d = −0.83, p = .002) with the corresponding CN-vs-MCI-AD contrast non-significant. One feature, early target fixation count ratio at baseline, separated CN participants from MCI-AD only. The subtype-specific effects were therefore concentrated in MCI-Other rather than MCI-AD, and the heterogeneous composition of MCI-Other constrains their interpretation.

In the subtype classification analyses, the CN-vs-MCI-AD classifier achieved an area under the curve of 0.850 (95% CI [0.765, 0.917]; n = 96), and the CN-vs-MCI-Other classifier achieved 0.892 (95% CI [0.816, 0.953]; n = 90). Both classifiers used age-, sex-, and education-residualized features, since splitting the MCI group breaks the pooled one-to-one match. MCI-Other was therefore directionally more separable from CN aging than MCI-AD, though the confidence intervals overlap substantially and this difference is suggestive rather than conclusive. The MCI-Other cell is the smallest and most heterogeneous, including DLB and PD cases whose disease-intrinsic oculomotor abnormalities could themselves contribute to discrimination.

## Discussion

Naturalistic visual search in MCI was disrupted across distractor conditions, with the largest group differences occurring under perceptually similar distractors and, for early target capture, in the no-distractor baseline, a pattern that was largely attenuated at the whole-task aggregate level. Across these conditions the group differences ran in a counterintuitive direction: participants with MCI engaged both the targets and distractors less than CN participants, rather than being more captured by the distractors, a direction more consistent with diminished structured search than with preserved distractor suppression. The prediction that disruption would be expressed condition-specifically rather than as a uniform decrement was therefore partially supported: group differences were present at every condition tested, but their magnitude did not order with distractor demand, and which measure carried the largest difference varied by condition. This condition-specific disruption, carried by the parametric trial structure and substantially reduced when gaze metrics were collapsed across distractor conditions, was complemented by multivariate discrimination of CN from MCI, obtained within a single propensity-matched cohort under internal cross-validation, and by the replication of seven of the fourteen univariate panel measures in the same direction in an independent, held-out generalization sample. In etiology-based subtype analyses, both MCI-AD and MCI-Other were discriminable from CN aging, and the features carrying subtype-specific separation were concentrated in MCI-Other, whose heterogeneous composition constrains interpretation. Taken together, these findings indicate that naturalistic visual search captures meaningful behavioral differences between CN and mildly impaired adults that support multivariate discrimination, and that preserving the parametric trial structure carrying distractor-condition information recovers condition-specific univariate effects that are largely attenuated when gaze metrics are collapsed to the aggregate level, where only trial-to-trial variability in area-of-interest coverage separated the groups after correction, while the multivariate signal drew on features extending well beyond the condition-resolved univariate measures. To our knowledge, this is the first study to evaluate multivariate gaze-based classification from a multi-distractor-type naturalistic protocol. The classification result establishes that gaze behavior recorded during naturalistic scene search carries information that separates CN from MCI adults, and identifies which aspects of search behavior carry it.

### Distractor Engagement as a Marker of Structured Search

Read as a failure of inhibitory control, the reduced distractor engagement in MCI is unexpected, since diminished control would be expected to increase capture by salient competing objects rather than reducing it [21]. The pattern is more coherently read, however, once distractor engagement in a naturalistic scene is understood not as an index of inhibitory failure but as a marker of intact, structure-driven exploration: in an unimpaired visual system, perceptually and semantically salient objects, including the task-defined distractors, reliably draw fixation so that engaging a distractor reflects the same organized, object-directed scanning that also carries the viewer to the target. Consistent with this reading, distractor engagement and target engagement moved together rather than in opposition, both lower in the MCI group, while the corresponding share of dwell shifted onto background and peripheral regions; the group difference is therefore better characterized as a global reduction in engagement with named scene content than as selective sparing of the distractor, and the lower distractor engagement in MCI is more plausibly a consequence of search that is less driven by scene structure overall than a sign of preserved distractor suppression [21,22].

Engaging a distractor in this paradigm often requires actively discriminating it from the target, a comparison that is most demanding when the competitor is perceptually similar; the largest distractor effect observed here fell under perceptually similar distractors, which suggests that cognitively normal participants foveated and revisited the similar distractor in the course of resolving it against the target, whereas the more diffuse and less discriminative search characteristic of the MCI group engaged that comparison less. Capture by a salient object also depends on rapid, stimulus-driven orienting, and attenuated bottom-up orienting in MCI would reduce the pull a distractor exerts and lower engagement with it without any accompanying gain in top-down control [21]. These accounts are not mutually exclusive, and the present cross-sectional data cannot adjudicate between them; a residual, non-mechanistic possibility is that the distractor measures are expressed as proportions of a named-region gaze budget, so that the shift of MCI fixation toward unnamed background necessarily depresses every fixation, distractor and target alike. On each of these readings, distractor engagement during naturalistic search may be better interpreted as a signature of organized, salience-sensitive exploration than as a measure of distractibility, and its reduction in MCI is continuous with the broader disorganization of search indexed by the entropy and coverage measures. Future study will incorporate engagement and fixation behavior as covariates, and engineer new features that account for these differences, to more reliably isolate the effect of MCI on visual search behavior.

### Contextualization within the Literature

Exploration in the MCI group was both narrower and less consistent. Lower scanpath entropy over named regions is consistent with the reduced exploratory breadth documented across eye-tracking paradigms in MCI [15,21]. Two further measures move in the opposite direction: transition-matrix entropy, indexing the predictability of region-to-region transitions, and trial-to-trial variability in area-of-interest coverage were both elevated in the MCI group. Taken together these indicate that MCI-associated search is not simply a scaled-down version of CN search but is distributed across fewer regions while being less consistent from trial to trial and less predictable in its transitions, a dissociation that a single aggregate exploration index would obscure.

The two entropy measures are not redundant despite this apparent tension: scanpath entropy summarizes the marginal distribution of fixations across named regions, while transition-matrix entropy conditions on the region currently fixated, so a participant can concentrate fixations in fewer named regions overall (lower scanpath entropy) while still moving unpredictably among the regions actually visited (higher transition-matrix entropy). The engagement pattern is one of reduced dwell on both target and distractor regions in the MCI group, with a correspondingly greater share of dwell falling on background and peripheral regions, indicating diminished engagement with task-relevant scene content rather than selective capture by competing objects; this bears on prior evidence concerning attentional allocation and disengagement in MCI and across the Alzheimer’s disease spectrum [21,22]. The present condition-structured results also contextualize the aggregate pattern observed here: at the aggregate level, mean-level exploration measures did not separate the groups after false discovery rate correction, converging with the result reported by Eraslan Boz and colleagues [17], who found no significant difference in aggregate scanning behavior between amnestic MCI and healthy controls during real-world scene viewing. Read alone, such aggregate results have framed naturalistic search as insensitive to the MCI stage; read against the present condition-structured analyses, they instead indicate that collapsing across qualitatively distinct distractor contexts substantially attenuates the MCI signal rather than demonstrating its absence, a reading reinforced here by the one aggregate measure that did survive correction, trial-to-trial variability in area-of-interest coverage.

Preserving the distractor-condition structure recovered significant condition-specific interactions that were not detectable in the aggregate analysis, addressing the methodological limitation that has constrained prior naturalistic work [15,17]. The divergence from Eraslan Boz et al. [17] could, however, also reflect differences in sample size, stimuli, the breadth of features tested, or analytic flexibility rather than the preservation of condition structure alone; consistent with this caution, the aggregate scanpath-entropy effect did not itself survive false discovery rate correction, so condition-structure preservation cannot be established as the single decisive factor. These findings thus extend the prior literature by demonstrating that gaze-based search deficits are detectable within a naturalistic paradigm at the MCI stage, and not only in more advanced disease. Both MCI subtypes were separable from cognitively normal aging, and the features that carried subtype-specific separation were concentrated in MCI-Other rather than MCI-AD. The paradigm therefore appears sensitive to cognitive impairment at the MCI stage generally rather than to an Alzheimer’s-specific attentional signature.

Because the subtype split rests on consensus rather than biomarker-confirmed amnestic phenotypes and involves small and, for MCI-Other, heterogeneous cells, this pattern is best read as a constraint on disease-specific interpretation rather than as a positive claim about non-amnestic mechanism.

### Interpretation and Mechanism

The discrimination achieved by the full-feature classifier, considered alongside the weak aggregate mean-level effects and the significant condition-structured interactions, is consistent with a discriminative signal; the univariate analyses, which isolate condition structure, further indicate that this signal emerges when trial structure spanning multiple distractor categories is preserved rather than collapsed, although the classifier itself drew on the full feature space and so does not by itself establish that condition structure is necessary for discrimination. This discrimination, however, was obtained at a specificity of only 0.516 at the Youden operating point (barely above chance for ruling out CN participants), so the full-feature classifier’s practical value lies chiefly in its sensitivity, with the 14-feature panel’s more balanced operating characteristics (sensitivity 0.968, specificity 0.758) offering a more informative picture of what condition-preserving features can achieve. The features contributing to this discrimination were drawn from the broad search-task-specific feature pool rather than restricted to the condition-resolved versions of the aggregate exploration measures; the discrimination therefore cannot by itself be attributed to the condition-specific structure of those measures, and it is the univariate analyses, rather than the classifier, that localize the recovered group differences to condition-specific rather than aggregate levels. No single gaze measure was sufficient for discrimination; discriminative information was instead carried jointly by a compact set of features spanning distractor engagement, saccadic dynamics, and the organization of transitions between scene regions, consistent with disruption that is expressed across several aspects of search behavior rather than in any one canonical metric. The result was obtained under repeated cross-validation, but discrimination varied appreciably across alternative propensity-matching draws, so the estimate reported here is specific to this matched sample rather than a stable property of the paradigm. External validation in an independent cohort is required before any clinical interpretation is warranted.

Search disruption was expressed in both MCI subtypes, and the features carrying subtype-specific separation were concentrated in MCI-Other; the paradigm therefore appears sensitive to the MCI signal broadly rather than to an Alzheimer’s-specific attentional profile [21]. Interpretation of the subtype profiles is necessarily bounded: MCI-Other is a heterogeneous grouping comprising MCI attributable to DLB, VCID, PD, and to mixed pathology, and a mechanistic account of the MCI-Other pattern is therefore not warranted on the present data. The divergent gaze metrics in the MCI-Other group, particularly the elevated saccadic direction reversals, may in part reflect the intrinsic oculomotor and visuospatial abnormalities associated with the DLB and PD cases that group contains; whether these divergent metrics index an MCI-associated search deficit or the disease-intrinsic oculomotor pathology cannot be adjudicated on the present data, since the individual etiology cells were too small to analyze separately. Naturalistic visual search captured MCI-associated gaze changes that supported above-chance discrimination, with a discriminative signal present for both subtypes; resolving the mechanistic basis of the subtype profiles will require biomarker-confirmed longitudinal follow-up.

Early target capture at baseline, evident before any competing distractors were introduced, was the one measure in the subtype panel that separated CN participants from MCI-AD specifically. This may reflect deficits in proactive attentional orientation, consistent with the episodic-memory-binding impairments characteristic of MCI-AD [23,24], although it stands against a broader subtype pattern in which most features either separated both subtypes or separated MCI-Other alone. Oculomotor correlates of impaired memory encoding during visual tasks have been reported in clinical populations [25], providing a plausible link between gaze behavior at encoding and memory-binding processes.

### Limitations

Several limitations qualify the interpretation of these findings. While propensity matching improved the demographic comparability of the two groups, it downsampled the CN group to 62 participants, reducing statistical power for the detection of secondary effects. Further, the cohort was recruited from a clinic-adjacent, community-based research setting, and the primary sample was demographically matched by design.

Discrimination was weaker in the held-out generalization sample, where age, sex, and education differ between groups and the MCI base rate is lower. Matching therefore isolates the search signal from demographic confounding at the cost of a sample that is not representative of the population in which such a probe would be deployed, and replication in independent, population-based samples is needed before these findings can be generalized to the broader at-risk population.

Participants with subjective cognitive concerns were excluded because their inclusion attenuated discrimination: the panel scored them as more impairment-like (median predicted probability 0.37 vs. 0.24; p = .047), and their specificity at the matched operating threshold was correspondingly lower (0.45 vs. 0.57), even though no individual feature distinguished the two control subgroups after correction. This pattern, a subtle multivariate shift toward the MCI gaze profile among individuals reporting subjective concerns, is consistent with subjective cognitive concern marking an at-risk intermediate state and is a candidate signal in its own right; characterizing it will require a dedicated, adequately powered design with longitudinal follow-up, which we intend to pursue in future work.

False discovery rate correction was applied within each dependent variable’s family of two to three pairwise contrasts, which provides limited protection against type I error inflation across the full set of analytic dependent variables, and the ANOVA p-values themselves are uncorrected. Further, splitting the MCI group breaks the one-to-one demographic match, so the subtype contrasts rely on covariate residualization rather than matching and are correspondingly less well controlled than the pooled comparison. Both subtype cells are small, and MCI-Other is heterogeneous by construction, so the shared-vs-specific feature structure reported here should be treated as provisional and requires replication in samples large enough to analyze each non-Alzheimer’s etiology separately.

### Future Directions

The first priority for future study is external validation in an independent, population-based cohort to establish whether and to what degree the discrimination observed here transfers beyond a single matched sample. A second priority is deeper biomarker characterization, including a within-MCI dose-response analysis against plasma pTau217 and separate etiology-specific analyses once the individual non-Alzheimer’s etiology cells reach adequate sample sizes to support them. A third priority is longitudinal extension: tracking search performance over time within the parent cohort would establish whether baseline search disruption predicts subsequent cognitive decline and conversion to dementia, and would move the paradigm from concurrent discrimination toward prospective prediction. Finally, we intend to examine cross-modality integration within the BACAN battery, combining the visual search signal with the other tasks of the parent cognitive battery to establish the composite classification accuracy achievable across modalities and whether each modality adds discriminative value beyond the others.

## Conclusions

Naturalistic visual search in MCI was disrupted in a condition-specific manner across qualitatively distinct distractor conditions. A full-feature multivariate gaze-based classifier discriminated CN from MCI adults under cross-validation, even though mean-level differences were substantially attenuated at the whole-task aggregate level. These findings are consistent with attentional and cognitive differences between CN and MCI being detectable in a naturalistic behavioral search task. Naturalistic visual search, and the gaze-based features that carry the condition-specific disruption signal, therefore merit evaluation as a candidate early-detection probe in longitudinal and biomarker-stratified cohort studies.

## Acknowledgements

The authors thank the dedicated research participants and their study partners, faculty, staff, postdoctoral fellows, and trainees of the Comprehensive Center for Brain Health at the University of Miami Miller School of Medicine.

## Funding

Work on this study was supported by grants from the National Institute on Aging (R01 AG071514, R01 AG069765, and R01 NS101483), the Alzheimer’s Association (AARF-22-923592), the Evelyn F. McKnight Brain Research Foundation, and the Harry T. Mangurian Foundation. The funders played no role in study design, data collection, analysis, decision to publish, or preparation of the manuscript.

## Author Contributions

MJK is responsible for conceptualization, writing the original manuscript, methodology, formal analysis, and data curation. MJK and JEG are responsible for funding acquisition. All authors contributed to reviewing and editing and approve the final manuscript.

## Conflict of Interest

Dr. Michael J. Kleiman is the founder and Chief Scientific Officer of SciKey, and receives consulting fees from Cognivue and Wellsaid.AI. Dr. Kleiman also owns intellectual property described in this manuscript. Dr. James E. Galvin is Chief Scientific Officer for Cognivue, and receives consulting fees.

## Data Availability

All data is available by request on our portal, https://data.umiamibrainhealth.org, following IRB approval and a Data Use Agreement with the University of Miami.

## Notes

### Competing Interest Statement

MJK is Chief Scientific Officer of SciKey. MJK and JEG receive consulting fees from Cognivue. MJK receives consulting fees from Wellsaid.AI.

### Author Declarations

IRB of University of Miami gave ethical approval for this work

